# COVID-19 contact tracing during first delta wave, Lebanon, 2021

**DOI:** 10.1101/2022.01.31.22268856

**Authors:** Maryo Baakliny, Julia Naim, Aya Hage, Tatiana Tohmeh, Reem Abdel Malak, Joelle Bassila, Hawraa Sweidan, Mohamad Zein, Darine Wehbeh, Kassem Chaalan, Lina Chaito, Nada Ghosn

## Abstract

**Background:** On the 11th March, the WHO has declared COVID-19 pandemic. Officially, the virus was introduced in Lebanon on the 21st February 2020. Since then, the national curve has drawn several waves. From July to September 2021, Lebanon has experienced the first delta wave. As part of the investigation, contact tracing was enhanced to limit virus transmission. The objective is to describe close tracing approach and profile of close contacts identified during the first delta wave.

**Methods:** COVID-19 surveillance is integrated within the national communicable diseases surveillance. The case definitions are adopted from WHO guides. Laboratories report positive cases on daily basis to the Ministry of Public Health, on DHIS2 platform directly or indirectly via excel files importation. Once reported, case investigation is initiated. It includes contact tracing with: 1) identification of close contacts, 2) advice on quarantine and self-monitoring, 3) contact testing. Referral to field testing is made available free of charge for close contacts. Collected data is updated on DHIS2 platform. Later, data is cleaned and analyzed to generate the daily report including description of close contacts. The report is shared with decision makers, professionals, media and public.

**Results:** From week 2021W27 to week 2021W40, 85490 cases were reported. Case investigation rate reached 78.8% of the cases were investigated within 24 hours. 66.5% of investigated cases shared lists of contacts, with 3.6 as average number of contacts per case.

We identified a total of 161805 close contacts, 95% were from family members, 71% were not vaccinated and 10 % had prior COVID-19 infection.

As for contact testing, 65% had RT-PCR test upon investigation, with 32% positive result. Furthermore, 19205 were referred and tested via field testing, with 25% positivity rate. Of all identified contacts, the reported positive tests reached 56,904 representing 35.2% of all contacts.

**Conclusion:** During community transmission and mitigation strategy, contact tracing contributes to increase awareness to the contacts and importance to abide to quarantine measures and thus to slow down the virus circulation.

Current close contacts are characterized with new profile of prior infection, vaccination history and testing behavior. There is need to adapt the quarantine measures to close contacts based on their profile, and to ensure easy access to free testing.

## Background and objective

The coronavirus disease COVID-19 has been considered the main public health threat throughout the past 100 years (1). Since the start of Pandemic and up to September 2021, more than 4.7 million related deaths were reported (2). The SARS-CoV2 delta variant, identified in December 2020, has become the main concern and main epidemic strain worldwide. This variant was considered as a Variant of Concern (VOC) (3) and was responsible for new waves of COVID-19 infections (emerging variants) as declared by the World Health Organization (WHO).

To limit COVID-19 person-to-person transmission and slow the spread of the virus, various control measures are indicated: raise awareness on healthy behavior, detecting cases, detecting contacts, isolation and quarantine, infection control practice in health facilities, social measures, vaccination… Despite being in in the context of community transmission, the contact tracing is still valid option to slow down the epidemic curve and reduce mortality burden (4, 5, 6).

The virus had the first detected case in Lebanon on the 21st of February 2020. Since then, the national curve has drawn several waves (7). Since July 2021, the country is experiencing waves due to the Delta variant (8). Integrated within the case investigation, contact tracing was enhanced as it contributes to limit the virus spread in a period where hospitals had limited resources due to national economic crisis, and vaccination levels were still below 50% (7).

The main objectives are: 1) to describe COVID-19 close contacts identified during the first delta wave, and 2) to assess contact tracing performance and outputs indicators.

## Methods

COVID-19 is an immediately notifiable disease by laboratories and hospitals from both public and private sectors. COVID-19 confirmed case is a patient with positive PCR and/or positive antigen rapid test (9).

Laboratories report positive cases on daily basis to the Epidemiological Surveillance Program at the Ministry of Public Health, on DHIS2 online platform directly or indirectly via excel files importation (10).

Once reported, case investigation is initiated. Investigators call the cases within 24 hours in order to: 1) Get information on demographic characteristics, clinical presentation, underlying conditions, case management, exposure, vaccination status, 2) Provide health education message on isolation, self-monitoring and referral to hotlines, 3) Initiate contact tracing. If the call is not answered, a SMS message is sent advising the patient to call the hotline.

Contact tracing includes the following: 1) Identification and listing close contacts, 2) Provide advice on quarantine and self-monitoring, 3) Refer to contact testing, 4) Call the contacts if symptomatic. Detailed Standard Operating Procedures were established to harmonize the team work.

Close contact is defined as anyone who had direct contact with, or was within 1 meter for at least 15 minutes, with a person who is infected with COVID-19 while infectious or potentially infectious, even if the person with the infection did not have symptoms (9, 11). Lists of close contacts with basic information (name, age, relationship, onset of illness, prior infection, vaccination status, testing result, phone number) are collected and entered on Google sheets form than uploaded on DHIS2 platform using tracker capture (10).

Identified close contacts are targeted for SMS message advising to call the COVID-19 hotline (24 hours, 7 days) to get advice on quarantine, self-monitoring and free testing.

During the case investigation, referral to field testing is proposed for the close contacts meeting the following criteria:

- Showing symptoms
- Or not tested yet, without symptoms or prior infection (in past 3 months), starting day 7

The field testing system is set by the MOPH to provide free access to the close contacts. It includes a weekly schedule of sites in almost all districts. The sites are selected in open air areas as public gardens, stadiums, parking areas, hospitals, and medical centers… At those sites, trained staff (from MOPH, NGO and municipalities) collects data and specimen (nasopharyngeal swab in viral transport media). Lists of referred close contacts to field testing are shared with the field teams. The specimens are referred to designated laboratories for RT-PCR testing: Lebanese University laboratories in Hadath and Tripoli, Baalbeck Governmental hospital and the National Influenza Center at Rafic Hariri University Hospital.

In addition, identified symptomatic contacts are targeted for secondary investigation. During the call, data is collected related to person characteristics (demographics, profession, exposure, and vaccination status), knowledge on prevention measures, illness, and available test result. Also, the contact is provided with test appointment if needed and advice.

All collected data is managed via the DHIS2 platform where 4 forms are set: case investigation filled online, contact investigation filled online, uploaded contact list, and uploaded field testing.

Contact tracing is monitored via a basket of indicators generated on daily and weekly basis reflecting the performance of the team and of contact tracing: total number of calls, completed case investigations, cases contacted within 24 hours of reporting, new contacts, percentage of cases who shared contacts, referrals for free testing, contacts tested at field sites, contacts with positive result, average of contacts per case, profile of close contacts (11, 12, 13).

The figures are included in the daily national COVID-19 surveillance report. The report is shared with decision makers, professionals, media and public, and posted on the MOPH website (7).

## Main Results

### 1) Epidemic curve

From week 2021W27 to week 2021W40, 85490 cases were reported. 96.7% were locally acquired, and 3.3% were travel related. The epidemic cure shows a new wave that started at week 27 and peaked at week 32, then decreased (Figure 1).

**Figure 1:**
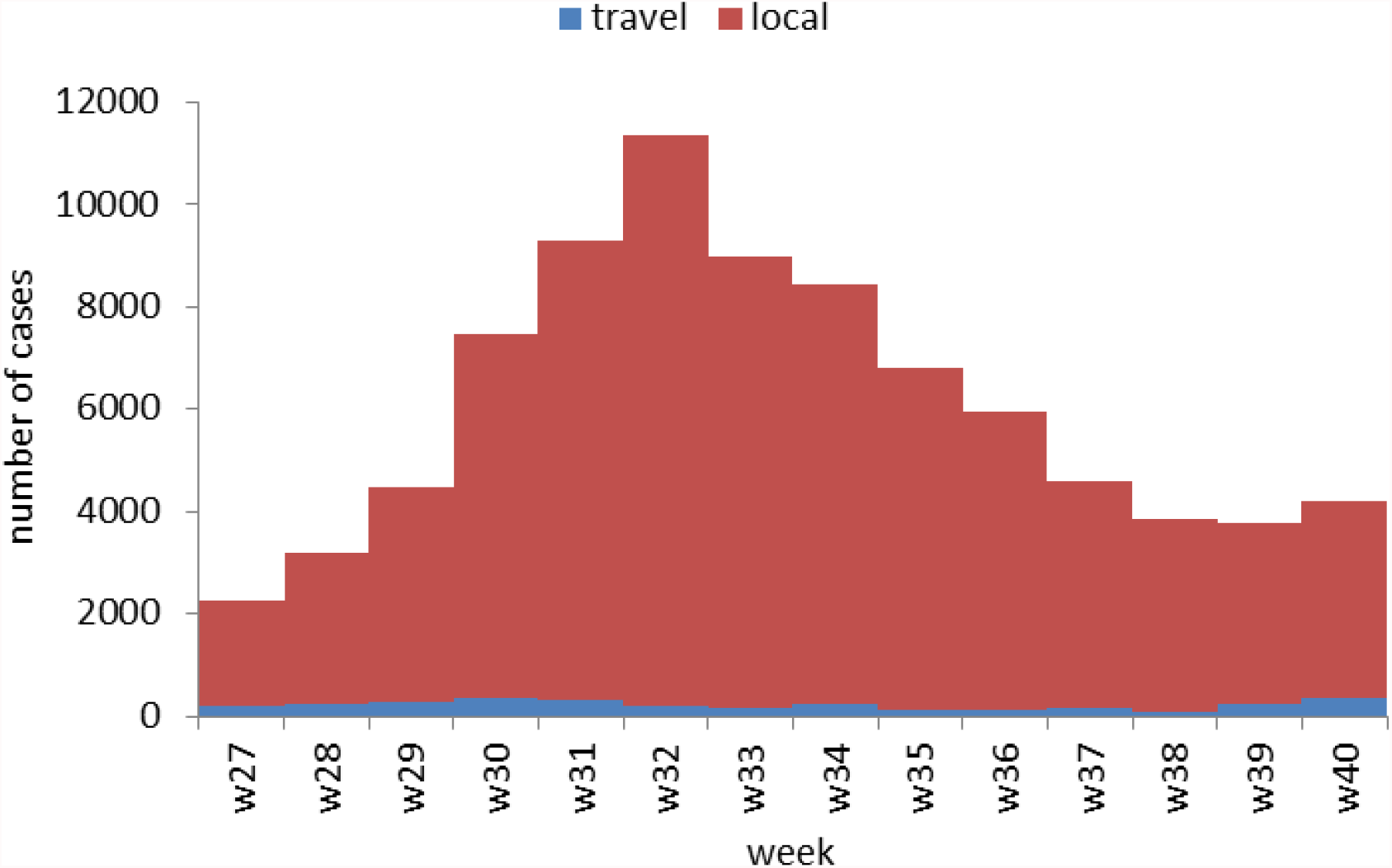
COVID-19 reported confirmed cases by week, Lebanon, 2021W27 – 2021W40

**Figure 2:**
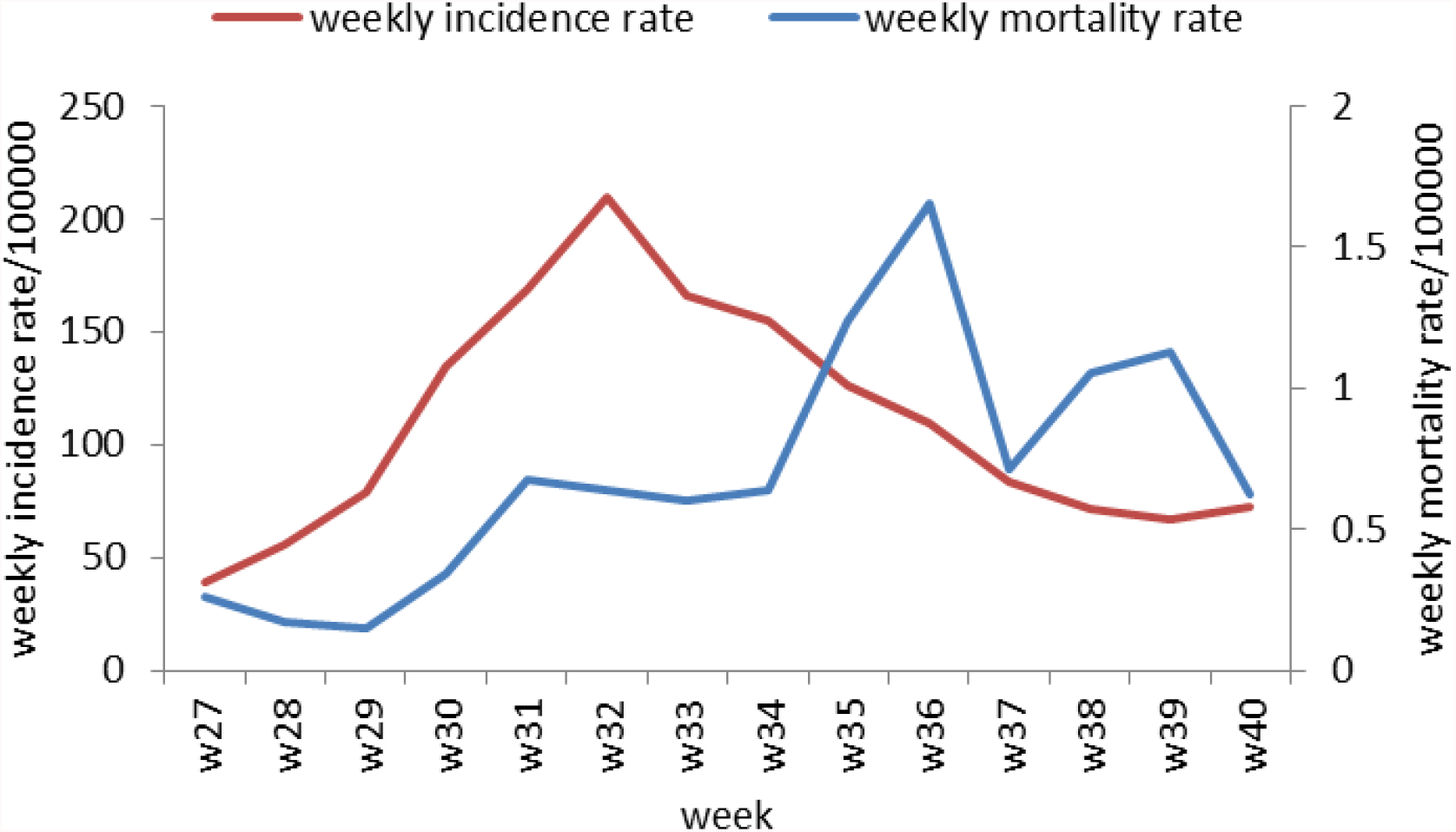
COVID-19 incidence and mortality rates per 100000 by week, Lebanon, 2021W27 – 2021W40

The weekly local incidence rate started at 38.6/100000 at week 2021W27 and reached 209.8/100000 at week 32 and decreased to 66.5/100000 at week 2021W39. As for the weekly mortality rate, the lowest was 0.17/100000 at week 2021W29, then increased to 0.68/100000 at week 2021W31 with some plateau, and increased again up to 1.65/100000 for week 2021W36, than raged from 0.62/100000 to 1.13/100000 for the next weeks (Figure 3)

**Figure 3:**
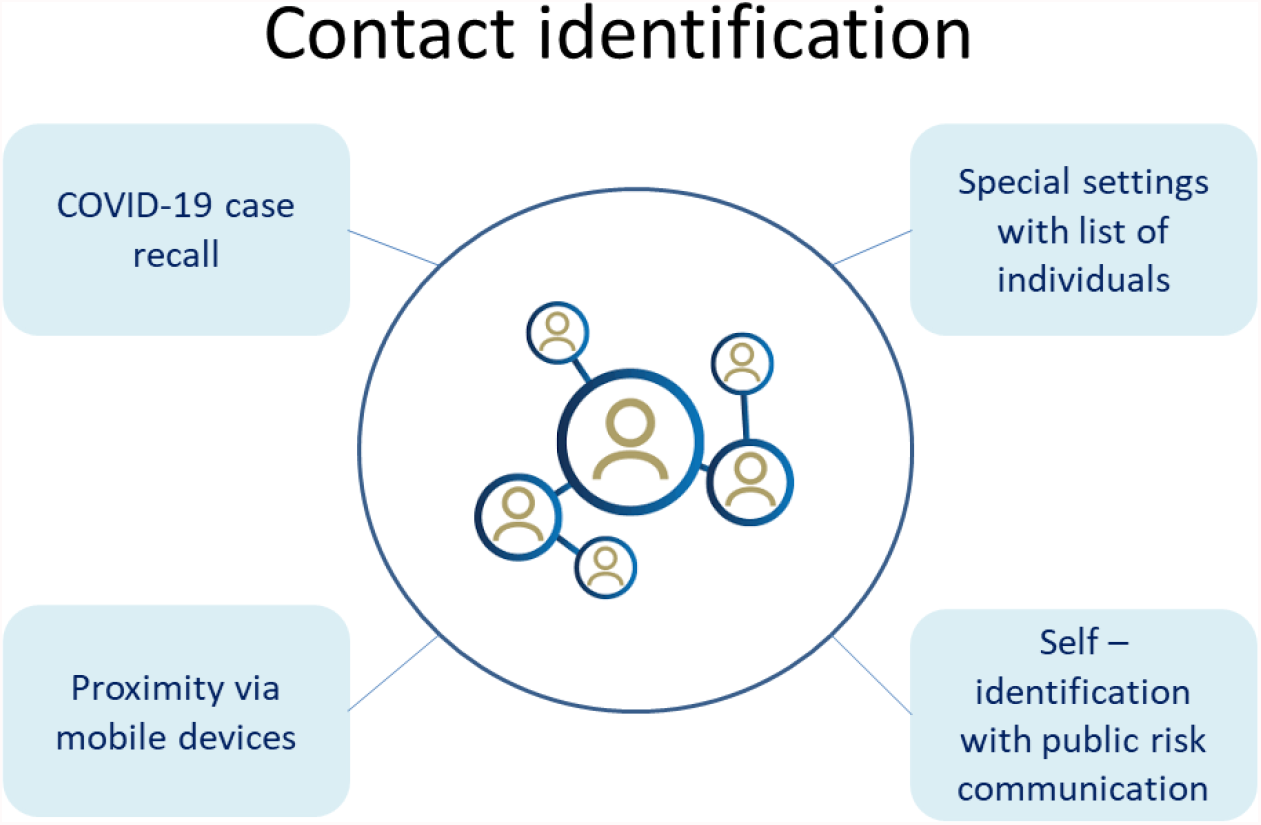
Mechanisms for close contacts identification

As per WHO classification, the community transmission level was level 2 on weeks 2021W27-28, reached level 3 for the weeks 2021W29-30, and level 4 for the weeks 2021W31-34, then returned to level 3 (15).

The wave is called first delta wave as genomic surveillance showed introduction and dominance of delta and delta variant (8, 16).

### 2) Case investigation

The 85490 new cases was the number after removing 7735 duplicates. The duplicate rate was 8.3%. Upon calling the cases: 90.1% were reached and agreed to be investigated, 0.3% reached and expressed refusal, and 9.6% were not reached. The causes for not reaching cases were: 55% wrong number, 35% no answer, 6% no phone number provided, and 4% international number provided (Table 1).

**Table 1:** Case investigation indicators

|  | N | % |
| --- | --- | --- |
| Reached and agreed | 77003 | 90.1% |
| Reached and refused | 239 | 0.3% |
| Not reached | 8248 | 9.6% |
| No answer [2] | 2654 | 35% |
| Wrong number [3] | 4157 | 55% |
| International number [4] | 266 | 4% |
| No phone number [6] | 446 | 6% |
|  | 7523 | 100% |
| Total | 85490 | 100.0% |

For cases with non-answer, 33420 SMS messages were sent advising for case isolation, safety measures, and seek advice from COVID-19 call center.

### 3) New contacts

A total number of 161,805 new contacts were identified (Table 2). The average age was, with age distribution: 37% aged 0-19y, 29% 20-39y, 24% aged 40-59y, and 9% 6-years and above. 52.2% of contacts were female and 47.8% male. Of all contacts, 95% were family relatives, 3% friends, and 1% co-workers. Of all contacts, 10% had prior infection (past 3 months). As per vaccination status, 22% had received 2 doses, 7% one dose and 71% were unvaccinated.

**Table 2:**
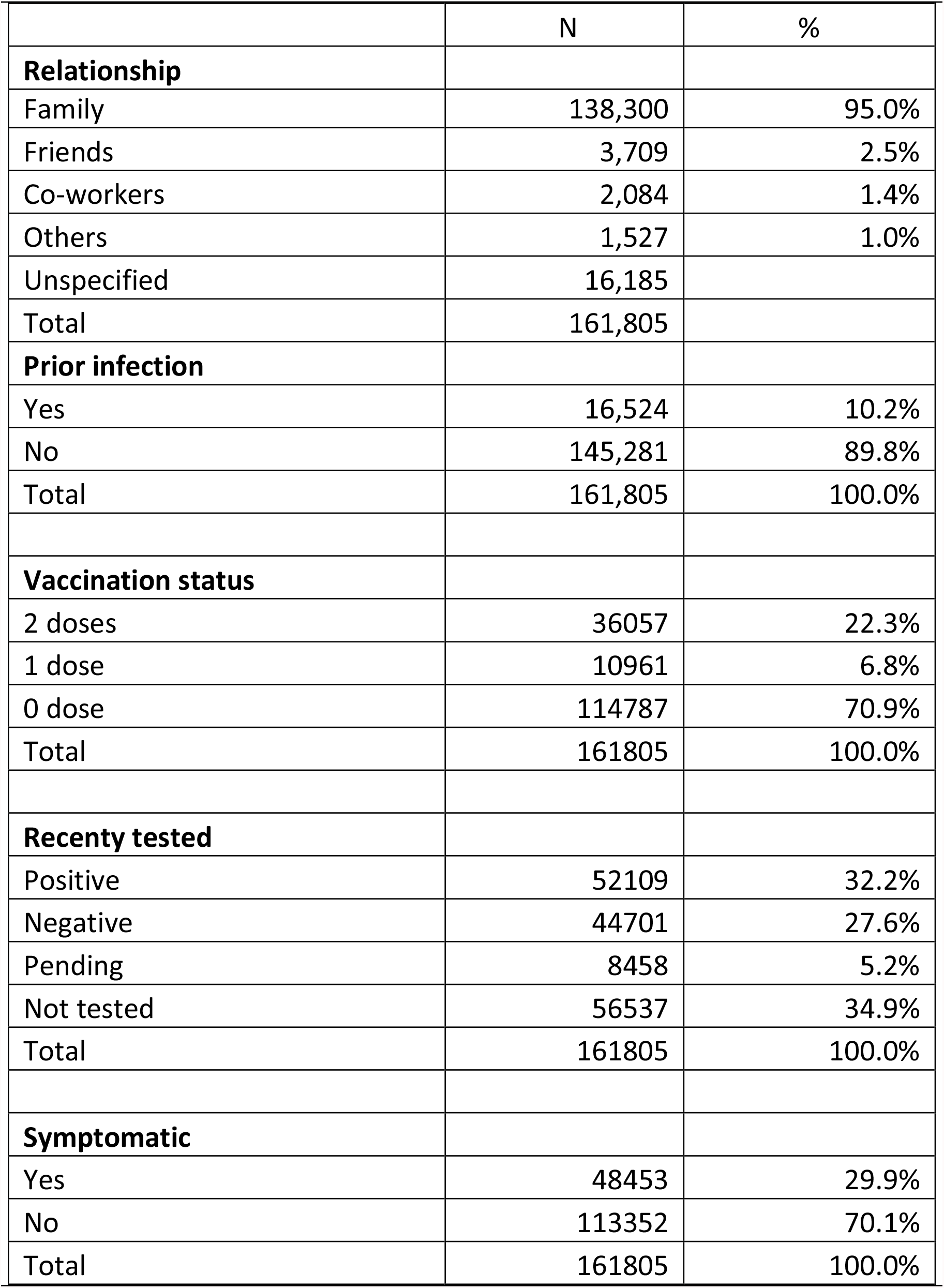
Characteristics of COVID-19 close contacts as reported by cases, Lebanon, 2021W27-2021W40

|  | N | % |
| --- | --- | --- |
| **Relationship** |  |  |
| Family | 138,300 | 95.0% |
| Friends | 3,709 | 2.5% |
| Co-workers | 2,084 | 1.4% |
| Others | 1,527 | 1.0% |
| Unspecified | 16,185 |  |
| Total | 161,805 |  |
| **Prior infection** |  |  |
| Yes | 16,524 | 10.2% |
| No | 145,281 | 89.8% |
| Total | 161,805 | 100.0% |
| **Vaccination status** |  |  |
| 2 doses | 36057 | 22.3% |
| 1 dose | 10961 | 6.8% |
| 0 dose | 114787 | 70.9% |
| Total | 161805 | 100.0% |
| **Recenty tested** |  |  |
| Positive | 52109 | 32.2% |
| Negative | 44701 | 27.6% |
| Pending | 8458 | 5.2% |
| Not tested | 56537 | 34.9% |
| Total | 161805 | 100.0% |
| **Symptomatic** |  |  |
| Yes | 48453 | 29.9% |
| No | 113352 | 70.1% |
| Total | 161805 | 100.0% |

At the time of investigation, 65% of all contacts had been tested with 32% positive test, 28% negative test and 5% with pending results. 35% were not tested yet. At the time of investigation, 30% of contacts were symptomatic.

In addition, 35,264 SMS messages were sent targeting 126,335 contacts with coverage of 78% (over all contacts). As most of contacts were family members sharing same mobile phone number, one SMS message was sent per phone number. The message recommends quarantine, contacting the hotline for advice and appointment for free testing.

### 4) Referral to COVID-19 testing

As part of contact tracing approach, the contacts who were not tested yet were invited to the nearest free testing sites supported by the MOPH. From week 27 to week 49, a total of 19,205 contacts were tested at field sites, with 25% positive result.

Of all identified contacts, the reported positive tests reached 56,904 representing 35.2% of all contacts.

### 5) Selective Contact investigation

Selective contact investigation was initiated starting week 2021W37 for those reported as symptomatic during case investigation.

For the weeks 2021W37-40, 687 contacts were investigated and reached. 53% were female. 59% were aged 20-59 years. 92% were family contacts. 7% had prior COVID-19 infection. 74% were unvaccinated. 31% reported having received SMS message. 7% had called the COVID-19 call center. 89% confirmed to be symptomatic. 76% were tested. 18% get appointment from field testing. Among available results, 77% were positive. 97% had advice and guidance on quarantine. 16% did not abide to quarantine measures, whereas 83% had at least 7 days of quarantine (Table 3).

**Table 3:** Characteristics of COVID-19 close contacts investigated, Lebanon, 2021W37-2021W40

|  | N | % |
| --- | --- | --- |
| **Age group** |  |  |
| 0-9 | 105 | 15% |
| 10-19 | 124 | 18% |
| 20-29 | 116 | 17% |
| 30-39 | 119 | 17% |
| 40-49 | 91 | 13% |
| 50-59 | 78 | 11% |
| 60-69 | 28 | 4% |
| 70-79 | 13 | 2% |
| 80+ | 13 | 2% |
| **Sex** |  |  |
| Female | 366 | 53% |
| Male | 321 | 47% |
| **Contact profile** |  |  |
| Family | 528 | 92% |
| Friends | 8 | 1% |
| Other | 13 | 2% |
| Workers | 24 | 4% |
| Unsp | 114 |  |
| **Prior covid infection** |  |  |
| No | 627 | 93% |
| Yes | 49 | 7% |
| Unsp | 11 |  |
| **Vaccination** |  |  |
| 0 dose | 478 | 74% |
| 1 dose | 32 | 5% |
| 2+ doses | 140 | 22% |
| Unsp | 37 |  |
| **Received SMS message** |  |  |
| No | 447 | 69% |
| Yes | 199 | 31% |
| Unsp | 41 |  |
| **Called Call Center** |  |  |
| No | 605 | 93% |
| Yes | 43 | 7% |
| Unsp | 39 |  |
| **Symptomatic** |  |  |
| No | 70 | 11% |
| Yes | 577 | 89% |
| Unsp | 40 |  |
| **Tested recently** |  |  |
| No | 166 | 24% |
| Yes | 517 | 76% |
| Unsp | 4 |  |
| **Appointment to field testing** |  |  |
| No | 534 | 82% |
| Yes | 117 | 18% |
| Unsp | 36 |  |
| **Available lab result** |  |  |
| Negative | 112 | 23% |
| Positive | 381 | 77% |
| **Guidelines for quarantine/isolation** |  |  |
| No | 17 | 3% |
| Yes | 564 | 97% |
| Unsp | 106 |  |
| **Duration of quarantine (days)** |  |  |
| 0 | 112 | 16% |
| 1-6 | 8 | 1% |
| 7-14 | 438 | 64% |
| 15+ | 129 | 19% |

### 6) Contact tracing indicators

The weekly monitoring of the indicators allows measuring the performance and improving it, and assessing the health response national capacity (11, 12, 13, 14).

The percentage of cases investigated was 89.4% at week 27 and improved up to 94.3% at week 40, with overall value of 83.1%. The percentage of cases investigated within 24 hours (of case notification), improved from 48.5% at week 27 to 82.5% at week 40, with overall value of 78.8%.

The percentage of cases who shared lists of contacts was 54.6% at week 35 an increased to 74.7% at week 40, with overall value of 66.5%.

As per average number of contacts per case, the overall value was 3.56 (Table 4).

**Table 4:** Contact tracing indicators, Lebanon, 2021W27-2021W40

| Indicator | W 27 | W 28 | W 29 | W 30 | W 31 | W 32 | W 33 | W 34 | W 35 | W 36 | W 37 | W 38 | W 39 | W 40 | W 27-40 |
| --- | --- | --- | --- | --- | --- | --- | --- | --- | --- | --- | --- | --- | --- | --- | --- |
| % of investigated cases | 89.4 | 87.6 | 85.8 | 87.4 | 88.1 | 90.6 | 92.3 | 93.7 | 94.2 | 93.9 | 94.1 | 94.4 | 93.5 | 94.3 | 83.1 |
| % of cases reached within 24 hours | 48.5 | 97.5 | 79.8 | 76.2 | 56.9 | 80.5 | 79.5 | 80.5 | 79.5 | 71.8 | 90.9 | 86.3 | 93.1 | 82.5 | 78.8 |
| % of cases who shared list of contacts |  |  |  |  |  |  |  |  | 54.9 | 63.9 | 59.3 | 68.3 | 67.1 | 74.7 | 66.5 |
| Average contacts per case | 3.79 | 3.84 | 3.82 | 3.56 | 3.53 | 3.38 | 3.52 | 3.19 | 3.52 | 4.04 | 3.9 | 3.8 | 3.83 | 4.06 | 3.56 |

## Discussion

### 1) Types of approaches for contact tracing

We can distinguish various strategies in contact tracing.

The objectives of the contact tracing differ between containment and mitigation phase. During containment, the objective is to contain the outbreak at the source and rigorous contact tracing is applied. During mitigation, the objective is to contribute with other measures to slow down the spread of the virus.

The identification of close contacts can be performed via 4 pathways: a) identification via the cases based on patient recall (systematic or selective case investigation); b) identification via proximity using mobile devices and GPS and Bluetooth technology; c) identification via special settings with documented lists of attending individuals (such in airplane, school, prison…); and d) self-identification (Figures 3, 4).

**Figure 4:**
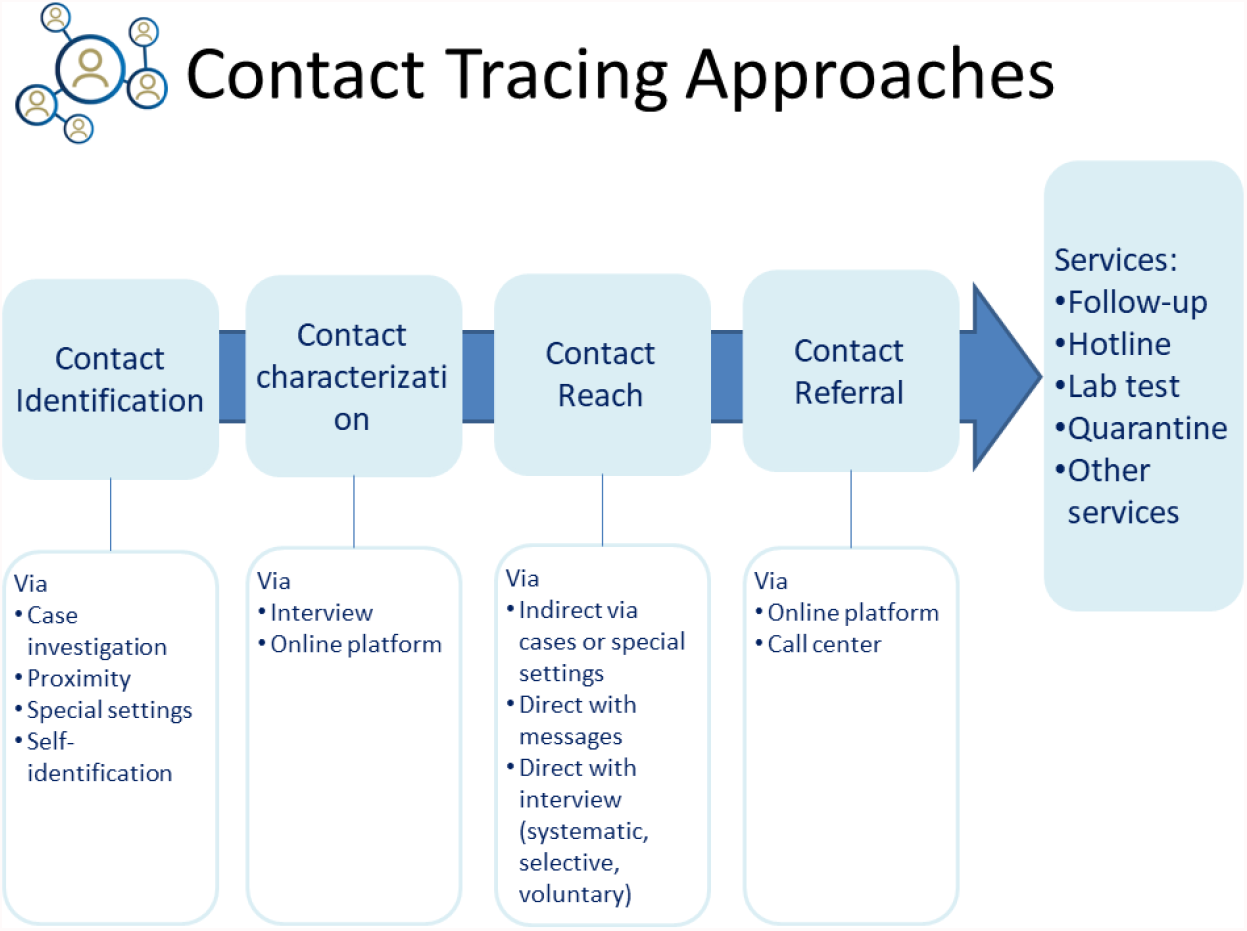
Contact tracing from identification to services pathway

The characterization of the contacts in term of exposure with the cases and identification of the high risk group can be done via: a) interview of cases and/or contacts and/or organizers; b) availability of online platform filled by cases, contacts, organizers, or contact tracing teams.

Reaching the contacts to provide them with advice on quarantine, self-monitoring and testing can be conducted via: a) Indirectly via the cases or the organizers, b) Directly via messages sent to the contacts (SMS, emails…), c) Directly with contact interviews that can be systematic (all contacts), selective (selected priority contacts), or voluntary (contacts calling dedicated hotline).

Additional attributes are linked to contact tracing:

- Follow up: it can be active follow-up with dedicated team calling the contacts using time-based protocol, or passive with dedicated online platform filled by the contacts
- Availability of hotline (24/7) to provide contacts with needed advice and available services
- Referral to laboratory free testing with specific protocols for symptomatic and asymptotic contacts
- Referral to quarantine services
- Documentation of the contact status for schools and workplaces …

As per the above typology, the contact tracing in Lebanon was: period 2021W27 to 2021W40:

- Identification and characterization via systematic case interview
- Reaching the contacts via SMS message, selective interview (for symptomatic starting week 2021W37) and voluntary interview via the hotline
- Referral to hotline, and free testing

The active follow up and referral to quarantine services were implemented at the early phase of the COVID-19 outbreak in Lebanon (Feb-Jul 2020) before reaching the community transmission.

### 2) Resources

Case investigation and contact tracing requires resources including: human resources, information and communication technology infrastructure, and referral pathways to health services.

The human resources are the critical point to decide on the adopted approach for contact tracing. At least, there is need to ensure enough staff to conduct the systematic case investigation. The needed capacity can be estimated based on population size (17, 18), or based on disease burden (19). An estimation of needed human resources based on population size for contact tracing with systematic contact interview and follow up, the needed capacity will increase to 15/100000 which will require 900 staff with adequate supervision and monitoring (17, 18). In Lebanon, with the support from the World Health Organization and the Lebanese Red Cross, the daily capacity included 60 agents, with maximal capacity of 4.9 calls/hour/investigator.

Moreover, the profile of the investigators affects the quality of data collection. For regular daily activity, the tasks are better performed with recruited public health workforce, trained on COVID-19 investigation and contact tracing. The workforce can be scaled up, for critical short period with high case load, by volunteers in particular medical or paramedical students (20).

The information and communication technology (ICT) includes the tools used to reach the cases and contacts and document the collected information. Here, the online platform and the call center technology with soft lines offer a better working environment for the various involved agents and actors.

As for the referral to health services, there is need to demonstrate to the cases and the contacts the added value of the contact tracing in term of individual benefits. Health services include access to reliable information, laboratory test, medical consultation, hospital admission, quarantine service (21).

### 3) Training

The team involved in contact tracing needs to have initial and continuous training. Indeed, they need to be trained on SOPs and to have regular update of their knowledge on the national situation and measures, in addition to the global situation. Training includes also the efficient use of ICT tools, the communication skills and the stress management (22, 23, 24).

### 4) Monitoring performance indicators

Moreover, the monitoring of key performance indicator is considered as vital component in controlling the effectiveness of case investigation and contact tracing. A daily monitoring of selected indicators is essential to track the team performance, per administrative level and per agent. This will aim to identify weaknesses and technical problems, and to demonstrate the improvement to reach the targets such as 80% of case investigation (15), the 70%-75% for cases with shared lists of contacts (contact elicitation success rate) (25, 27), and 5 as average number of contacts per case (27).

The indicators vary across countries and by time. In similar analysis, in the USA, in June-July 2020, 57% of cases were interviewed, 1.15 was the median of contacts per case (28). The percentage of cases sharing lists of contacts ranged from 27% in Jun-Jul 2020 up to 45% in Oct 2020, and the average of contacts per case reached 2.6 to 3 in Washington State (29, 30). In USA Connecticut, the percentage of contacts notifiers reached 28% (31).

### 5) Public awareness

At community level, the public awareness on disease and contact tracing contributes to have better understanding and adherence of the cases and their contacts to the contact tracing measures: isolation, quarantine, sharing lists of close contacts, testing (32)… When initiating contact tracing, there is need to ensure adequate risk communication for the public, including statement related to the confidentiality. Confidentiality is among the frequent concerns of the citizens to provide adequate data (21).

On the other hand, the contact tracing contributes to deliver individual preventive measures, and to address the concerns of cases and controls, and to convince them to consider seriously adopting healthy behavior.

## Conclusion & Recommendations

Contact tracing is a component for optimal COVID-19 response, complementing public health measures: case detection, case isolation, vaccination, increase awareness.

The effectiveness of contact tracing depends on a number of factors including timely case investigation, timely identification of contacts, timely advice on self-monitoring, quarantine and access to laboratory testing. Additional factors are the community awareness, and the level of trust in public health authority and policies. The most important daily challenge resides in convincing individuals, in particular asymptomatic, for self-quarantine, self-monitoring and testing. As convincing tools, there is need to provide contacts with access to hotline, free testing, awareness material available online.

The contact tracing needs investment in resources including securing minimal human resources, and informatics and communication infrastructure. The available resources will determine the affordable approach for the country.

The analysis of contact profile in terms of spatial-temporal exposure to cases, prior COVID-19 infection, vaccination status, adherence to protective equipment and behavior identifies more and more contacts with prior infection, or with full vaccination status, and thus needing adapted recommendations for quarantine.

Finally, the contact tracing during community transmission and in the context of mitigation strategy and vaccination campaigns needs to have clear mandate, and to be assessed in terms of disease control.

## Data Availability

All data produced in the present study are available upon reasonable request to the authors

## Acknowledgment

We thank the World Health Organization to support the contact tracing at the Lebanese MOPH. We thank also the laboratories and the NGOs involved in field testing: Laboratory of Molecular Biology and Cancer Immunology at the Faculty of Sciences at the Lebanese University at Hadath, Health and Environment Microbiology laboratory at the Faculty of Public Health/ Doctoral School of Science and Technology at the Lebanese University at Tripoli, National Influenza Center at Rafic Hariri Univesrity Hospital, Baalbeck Governmental Hospital, United Nations High Commissioner for Refugees (UNHCR), Amel Association, International Orthodox Christian Charities (IOCC), Medecins Sans Frontieres (MSF), Armenian Cross, Islamic Health Society.

